# The yield on re-interpretation of genetic variants in pediatric cardiomyopathy

**DOI:** 10.1101/2024.12.20.24319248

**Authors:** Takanori Suzuki, Robert Lesurf, Rajadurai Akilen, Xiaoqiao Xu, Eva Maria Batke, Vinay J Rao, Rebekah Jobling, Laura Zahavich, Natasha Henden, Jodie Ingles, Edgardo Alania Torres, Keichi Hirono, Nathalie Roux-buisson, Perundurai S Dhandapany, Christoph Sandmann, Seema Mital

## Abstract

**Background:** Variant interpretation can change over time as new knowledge emerges. Our aim was to determine the frequency and causes of variant reinterpretation on systematic re-evaluation in pediatric patients with cardiomyopathy.

**Methods:** Overall, 227 unrelated pediatric patients with cardiomyopathy enrolled in the Heart Centre Biobank harbored a pathogenic/likely pathogenic (P/LP) variant and/or a variant of uncertain significance (VUS) on clinical genetic testing (2005-2022). Variant pathogenicity was re-evaluated using the American College of Medical Genetics and Genomics (ACMG) guidelines. Additional extension cohorts (n=4547, cases) were analyzed to assess variant burden in cases versus controls (gnomAD 4.1.0).

**Results:** 382 variants (110 P/LP, 272 VUS) in 227 patients were re-evaluated. Forty-nine variants in 49 patients (21.6%) changed classification. Twelve (10.9%) P/LP variants were downgraded to VUS in 14 patients. Leading criteria were high population allele frequency and variant not located in mutational hotspot or critical functional gene domain. Thirty-seven (13.6%) VUS were upgraded to P/LP in 35 patients. Leading criteria were variant location in mutational hotspot for gene, and deleteriousness on in silico prediction. Only 8 reclassified variants had been reported back by the clinical genetic testing laboratory at the time of the study. Ten of the 37 VUS upgraded to P/LP were significantly enriched in cardiomyopathy cases (n=4796) versus controls.

**Conclusions:** One in five patients with cardiomyopathy had a clinically relevant change in variant pathogenicity on systematic re-evaluation that would require modifying family clinical screening and cascade genetic testing. These findings underscore the clinical importance of regular variant re-interpretation on follow-up.

## INTRODUCTION

Cardiomyopathy is primarily a genetic disorder and a common cause of childhood heart failure [1]. Cardiomyopathy phenotypes include arrhythmogenic cardiomyopathy (ACM), dilated cardiomyopathy (DCM), hypertrophic cardiomyopathy (HCM), restrictive cardiomyopathy (RCM), and left ventricular non-compaction cardiomyopathy (LVNC). Dozens of genes are implicated in causing the disease, with most genotype-positive cases being autosomal dominant and caused by a rare DNA variant [1], and with considerable overlap in the genes associated with different cardiomyopathy subtypes [1, 2].

The genotype of a patient informs cascade clinical and genetic screening of first-degree relatives. Genotype-positive patients i.e., patient harbors a pathogenic/likely pathogenic (P/LP) variant in a cardiomyopathy gene, especially those with P/LP variants in sarcomeric and other cardiomyopathy-associated genes, tend to have a worse prognosis and their genotype may offer clinically meaningful insights for risk stratification and treatment decisions [3]. Importantly, when a patient is genotype-positive, family members are offered cascade testing for the variant, and only genotype-positive family members require ongoing echocardiographic screening for cardiomyopathy [4]. When a patient is genotype-negative i.e., does not harbor a P/LP variant, then all family members are recommended serial echocardiographic screening [4]. A previous study in HCM reported that, among 1,361 probands with non-benign variants, 917 (67.3%) harbored P/LP variants and 444 (32.6%) harbored only variants of uncertain significance (VUS), the latter reflecting incomplete or conflicting evidence for pathogenicity [5]. However, variant interpretation can change over time as new knowledge, additional functional validation data, and more accurate computational prediction tools emerge. A change in variant interpretation can change family screening recommendations. The 2024 American Heart Association & American College of Cardiology guidelines for HCM and the European Society of Cardiology guidelines for cardiomyopathies emphasize the importance of periodic variant re-evaluation [6, 7]. To improve the efficiency of variant re-evaluation, clinicians are advised to document the date when the genetic test results were initially assessed and to consider any new medical and scientific information that has become available since that time [8]. Small pediatric cardiomyopathy cohort studies have reported variant reclassification rates ranging from 8.9%-10.3% [9, 10]. Here we performed a real-world systematic reevaluation of variants in a larger cohort of unrelated pediatric cardiomyopathy cases to identify the frequency and criteria for clinically ‘actionable’ variant reclassification on follow-up.

## METHODS

### Study cohort

The study cohort included 227 unrelated patients with primary cardiomyopathy <18 years old, enrolled through the Heart Centre Biobank Registry at the Hospital for Sick Children (Toronto, Ontario, Canada) (**Table 1 and Supplementary Table S1**). Patients who harbored a P/LP variant or a VUS on clinical genetic testing were included. Patients with secondary cardiomyopathy caused by chromosomal malformations, neuromuscular disorders, mitochondrial or metabolic disorders, congenital heart defects, or reversible causes were excluded. Patient demographics, diagnosis, type of clinical testing (panel, exome), year of clinical genetic testing, the original (and updated) variant classification by the genetic testing laboratory were captured. In addition, to perform case control burden analysis for relevant reclassified variants, data was accessed from an extension cohort of 4547 cases derived from international studies and published literature (**Supplementary Table S2**). The gnomAD (v4.1.0 genome dataset) was used to assess allele frequencies (n= 76,215) for variant re-interpretation, but for case-control burden analysis, we used gnomAD (v4.1.0, exome dataset) (n= 730,947) to avoid any bias related to using overlapping patients as controls. The study was approved by the Institutional Research Ethics Boards and written informed consent was obtained from all patients and/or their parents/legal guardians and the study protocol adhered to the Declaration of Helsinki.

**Table 1.** Baseline study cohort characteristics.

| **Characteristics** | **N=227 (%)** |
| --- | --- |
| Male | 133 (58.6%) |
| **Ancestry** |  |
| European | 140 (61.7%) |
| Asian/South Asian | 56 (24.7%) |
| African | 22 (9.7%) |
| Mixed | 5 (2.2%) |
| Unknown | 4 (1.8%) |
| **Cardiomyopathy phenotype** |  |
| DCM | 104 (45.8%) |
| HCM | 76 (33.5%) |
| LVNC | 20 (8.8%) |
| RCM | 10 (4.4%) |
| ACM | 9 (4.0%) |
| Mixed type | 8 (3.5%) |
| **Type of test** |  |
| DCM panel | 25 (11.0%) |
| HCM panel | 54 (23.8%) |
| Multiple panels | 124 (54.6%) |
| Whole exome sequencing | 4 (1.8%) |
| Others | 3 (1.3%) |
| Unknown | 17 (7.5%) |
| **Variants identified** |  |
| P/LP | 110 (28.8%) |
| VUS | 272 (71.2%) |
| **Patient genotype** |  |
| Harbored P/LP variant | 71 (31.2%) |
| Harbored VUS | 122 (53.7%) |
| Harbored P/LP variant and VUS | 34 (15.0%) |

*ACM; arrhythmogenic cardiomyopathy, DCM; dilated cardiomyopathy, HCM; hypertrophic cardiomyopathy, LVNC; left ventricular non-compaction, RCM; restrictive cardiomyopathy, VUS; variant of uncertain significance.*

### Re-evaluation of variant pathogenicity

Variants identified as P/LP or VUS on clinical genetic testing were reinterpreted for pathogenicity using the American College of Medical Genetics and Genomics/Association for Molecular Pathology (ACMG/AMP) guidelines for interpretation of variant pathogenicity. Using these criteria, a variant can be interpreted as pathogenic, likely pathogenic (P/LP), VUS, likely benign (LB), or benign (B) [11, 12]. For variants with multiple isoforms, the Matched Annotation from NCBI and EMBL-EBI transcript (MANE Select and MANE Plus Clinical) was used to annotate and evaluate variant effects in standardized and clinically relevant gene transcripts. A clinically actionable variant reclassification was defined as either the upgrading of a VUS to P/LP, or the downgrading of a P/LP variant to a non-pathogenic variant i.e., VUS or B/LB variant.

Several criteria were applied for variant re-interpretation including confirmation of patient phenotype, the current association between gene and cardiomyopathy phenotype on the most recent ClinGen version 11 curation [13], updated ClinVar records (2024-01-29 Web Release), literature evidence, control population allele frequencies, and prediction scores derived from computational tools. When available, the results of clinical genetic tests from parents or siblings were utilized to ascertain *de novo* status and to perform segregation analysis for a familial variant. Reclassifications were verified by our Return of Research Results committee comprising pediatric cardiologists, clinical geneticists, genetic counselors, and bioinformaticians [14].

A detailed approach to variant reinterpretation is described below. Of note, all eligible variants reported on clinical testing were single nucleotide variants and small insertions-deletions.

1. PVS1 criterion was applied to loss-of-function variants in haploinsufficiency-intolerant genes using gnomAD 4.1.0, defined as a probability of loss-of-function intolerant (pLI) score ≥0.9. PVS1 criterion was not applied to *MYH7* variants as per ClinGen guidelines because the contribution of loss-of-function variants in this gene to inherited cardiomyopathy remains incompletely understood [15]. *TTN* variants were only considered pathogenic if they were protein-truncating variants (frameshift, nonsense, canonical splice site) predominantly in the A-band and I-band regions and isoforms [16, 17].
2. The PS2 criterion was applied when the variant was confirmed through genetic testing to be *de novo* with parentage confirmed and no family history of cardiomyopathy.
3. The PS4 criterion of higher variant burden in cases compared to controls i.e., gnomAD (v4.1.0, exome dataset; n= 730,947) was conducted by jointly analyzing cases from our cohort and additional cases from extension cohorts (n = 4,796).
4. The PM1 criterion was applied if the variant was located in a mutational hotspot or critical functional domain for the gene. This was determined using DiscoVari, the Cardiomyopathy Variant Curation Expert Panel from ClinGen, and/or literature evidence [18, 19]. For *MYH7*, the relevant transcripts analyzed were ENST00000355349 and NM_000257.4, covering codons 167 to 931. In the case of *MYBPC3*, the analysis focused on transcripts ENST00000545968 and NM_000256.3, specifically targeting codons 485 to 502 and 1248 to 1266. For *TNNI3*, the key transcripts used were ENST00000344887 and NM_000363.5, covering codons 141 to 209. For *TNNT2*, the transcripts ENST00000367318 (codons 79 to 179), and ENST00000656932.1/NM_001276345.2 (codons 89 to 189) were included in the analysis.
5. PM2 criterion was applied if population variant allele frequency in gnomAD 4.1.0 (genome dataset) was <0.0001 for autosomal dominant and <0.01 for recessive inheritance, while accounting for Grpmax filtering allele frequencies (95% confidence) as ancestry-specific allele frequencies [20].
6. The PM5 criterion was applied when the variant incorporated a novel missense change at an amino acid residue where a different missense change is determined to be pathogenic in ClinVar or Leiden Open Variation Database v.3.0 supported by literature [12, 21].
7. The PP1 criterion was applied when the variant segregated with disease amongst family members. According to the ClinGen guidelines, in cardiomyopathies with reduced penetrance, PP1 is applied depending on the number of affected individuals demonstrating co-segregation with the variant (not including the proband). Specifically, PP1 supporting is applied when co-segregation is observed in a single affected family member, PP1 moderate for 2-3 affected members, and PP1 strong for ≥4 affected members [22].
8. The PP2 criterion was defined as a missense variant in a gene with a low rate of benign missense variation, where missense variants are a known mechanism of disease. Based on ClinGen Cardiomyopathy Expert Panel Specifications to the ACMG/AMP Variant Interpretation Guidelines in the Criteria Specification Registry for cardiomyopathy [19], PP2 was applied only to variants in the gene *TPM1*.
9. The PP3 criterion for missense variants which requires multiple lines of computational evidence supporting a deleterious effect on the gene was applied when all three pathogenicity scores (CADD v1.7, REVEL, and AlphaMissense.v2023.hg38) reached pre-defined thresholds [23–25] These thresholds were adopted from the ClinGen recommendations for CADD (>25.3) and REVEL (>0.644) [26]. As the threshold for AlphaMissense was not specified in this recommendation, it was derived from the original publication as a score >0.564 [24].

### Statistical Analysis

Frequency of variant reclassification was compared using Fisher exact test by year of clinical testing (before vs. after 2015 i.e., publication of ACMG guidelines), cardiomyopathy phenotype (HCM vs. DCM), ancestry (European vs. non-European), sex (male vs. female) and type of genetic test (single gene testing/single targeted panel vs. multiple panels/exome sequencing). Patients with LVNC, ACM, or RCM were excluded from subgroup analysis due to limited sample size. The proportion of variants reclassified from VUS to P/LP and from P/LP to VUS was also compared using Fisher exact test. A p-value <0.05 was considered significant. Burden analysis was performed by comparing the frequency of each variant in cases versus controls, with allele count and allele number extracted for each variant from gnomAD v4.1.0 exome dataset. Fisher exact test was used to assess the difference in allele frequencies between cases and the gnomAD v4.1.0 exome dataset used as controls. Following the guidelines of the Cardiomyopathy Variant Curation Expert Panel from ClinGen, the strength of evidence was classified based on the lower bound of the 95% confidence interval (CI) for the odds ratio (OR), with strong evidence requiring a lower bound of ≥20, moderate evidence requiring ≥10, and supporting evidence requiring ≥5, and anything below 5 considered not met.

## RESULTS

### Study cohort characteristics

The baseline characteristics of the study cohort (n=227) are summarized in **Table 1**. Among the participants, 133 (59%) were male; 64% were of European descent, 25% were of Asian descent, 10% were of African descent, and 1% were of mixed descent or other ancestries. Of 227 participants, 203 underwent one or more gene panel testing, 4 underwent whole exome sequencing (WES), and 20 underwent other types of genetic tests. The 227 patients harbored a total of 382 variants (110 P/LP, 272 VUS) (**Supplementary Table S1**). 105 participants (46.3%) harbored at least one P/LP variant, 122 (53.7%) harbored only VUS, and 34 (15%) harbored a combination of P/LP variants and VUS. Three variants that were found in two families each - *MYBPC3* (c.442G>A, p.Gly148Arg), *MYH7* (c.1759G>A, p.Asp587Asn) and *LMNA* (c.868G>A, p.Glu290Lys) were reported differently for each family by the clinical laboratory i.e., P/LP for one family and VUS in another family.

### Reinterpretation of variant pathogenicity

#### Reclassification of P/LP variants

After reinterpretation, 12 of 110 P/LP (10.9%) variants were downgraded to VUS in 14 of 227 patients (6.2%) **(Table 2)**. Genes affected by the downgrade included *MYH7* (5 variants), *MYBPC3* (4 variants), and one variant each in *LMNA*, *DSG2*, and *RRAGD*. Notably, the *MYBPC3* c.3628-41_3628-17del (Δ25bp) variant was identified in six South Asian cases. The criteria primarily contributing to downgrading of P/LP variants were an allele frequency higher than expected in gnomAD (n=7), the variant not being in a mutational hotspot or critical functional gene domain (n=5), the variant not being predicted as deleterious by *in silico* tools (n=4), the variant recently reported as VUS in ClinVar (n=3), and failure to meet *MYH7*-specific ACMG/AMP criteria (n=1) (**Figure 1a**).

**Figure 1.**
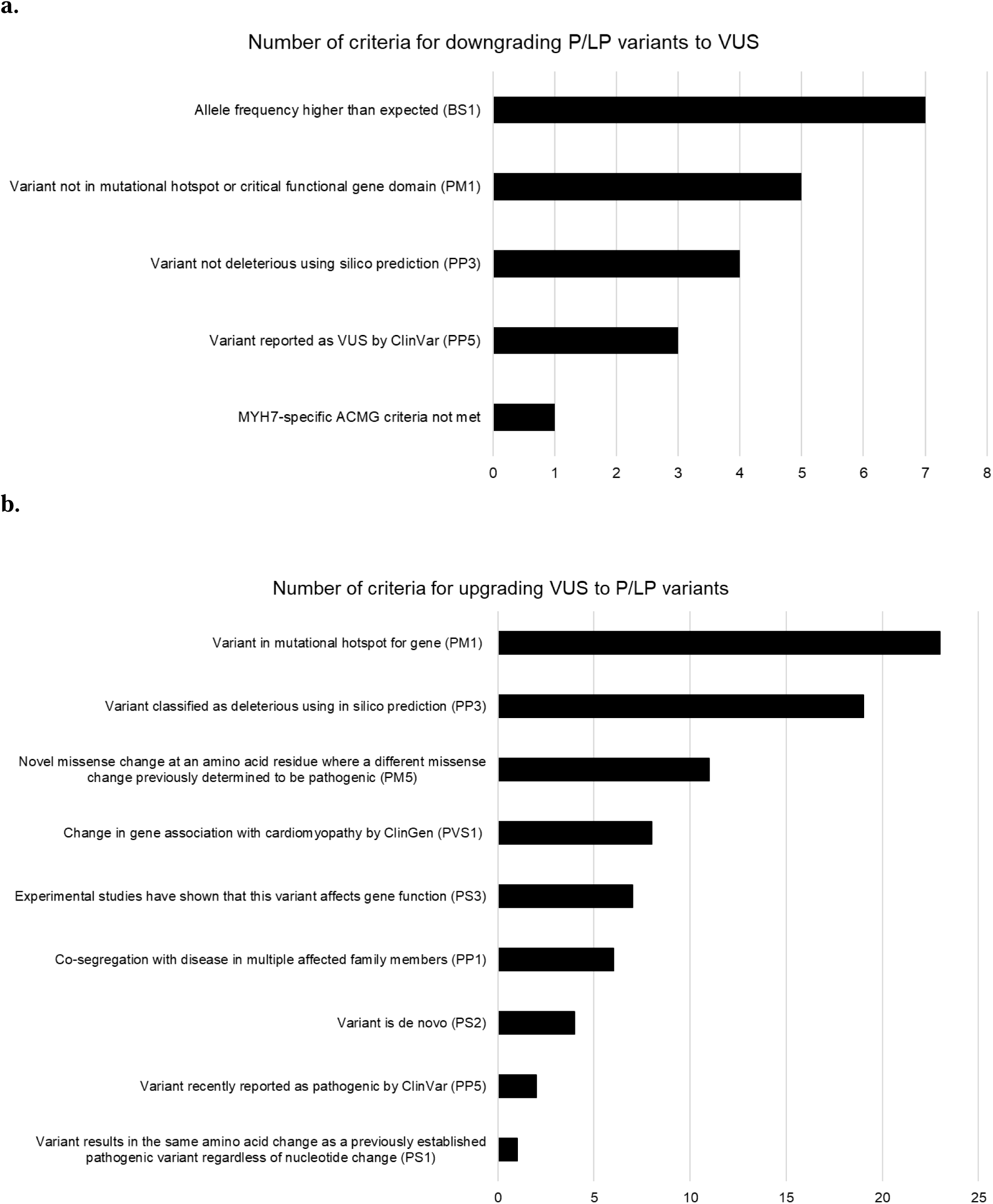
Criteria for variant reclassification. **(a)** Downgrading P/LP variants to VUS, **(b)** Upgrading VUS to P/LP variants. ACMG, American College of Medical Genetics and Genomics; P/LP, Pathogenic/Likely pathogenic; VUS, variants of uncertain significance.

**Table 2.**
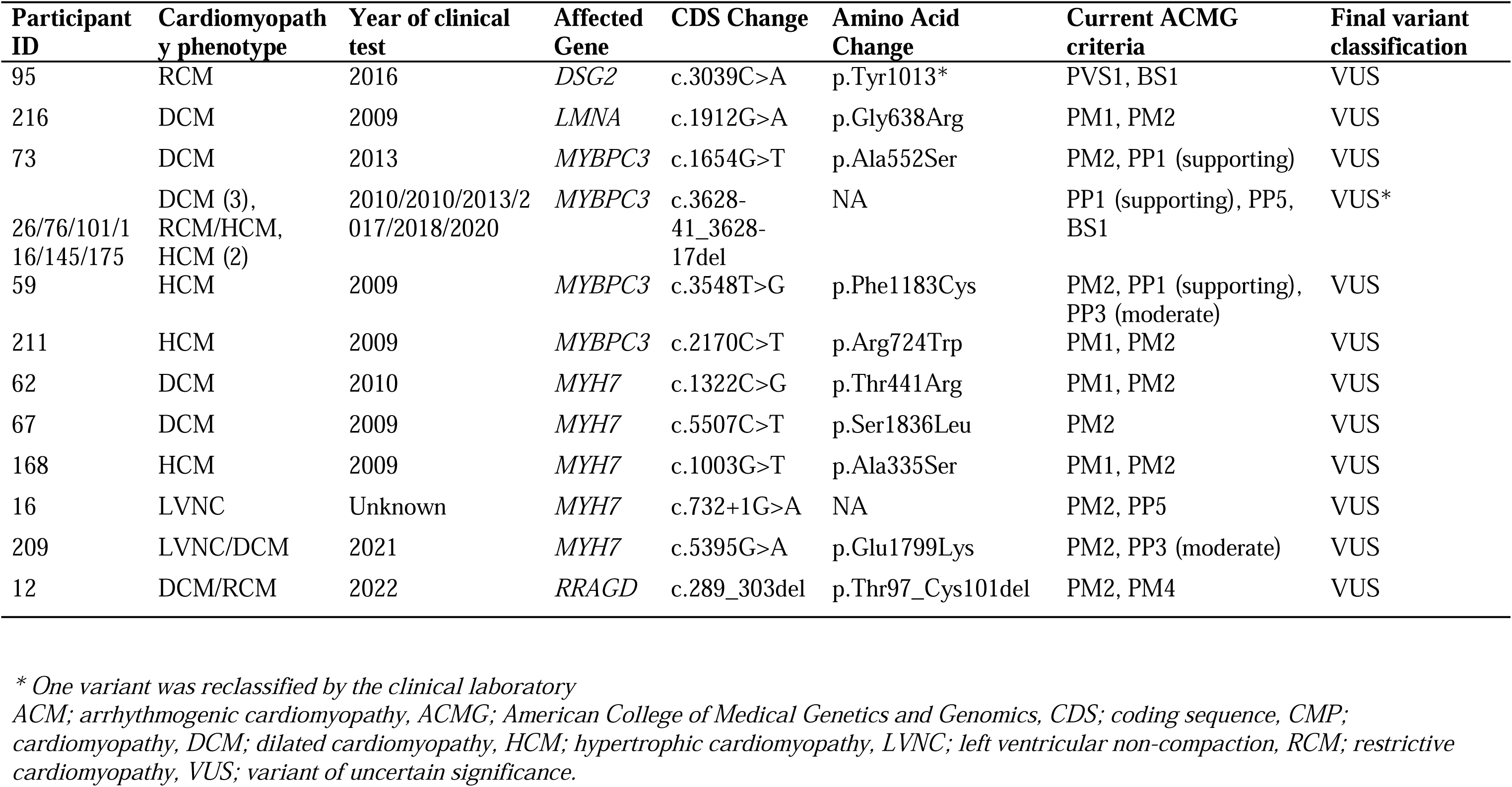
Pathogenic or likely pathogenic variants reclassified to VUS.

#### Reclassification of VUS

37 of 272 VUS (13.6%) were upgraded to P/LP in 35 of 227 patients (15.4%) (**Table 3**). Genes affected by the upgrade included *ACTC1* (1 variant), *ACTN2* (2 variants), *LMNA* (1 variant), *MYBPC3* (2 variants), *MYH7* (13 variants), *NEXN* (3 variants), *PLN* (1 variant), *TNNC1* (1 variant), *TNNI3* (2 variants), *TNNT2* (4 variants), *TPM1* (3 variants), *TTN* (1 variant), and *VCL* (2 variants). The primary criteria contributing to upgrading of VUS to P/LP were variant location in a mutational hotspot for the gene (n=23), variant interpretation as deleterious by modern *in silico* prediction tools (n=19), a novel missense change occurring at an amino acid residue where a different pathogenic missense change had been observed before (n=11), a change in the gene association with cardiomyopathy according to ClinGen (n=8), published experimental findings suggesting that the variant affects gene function (n=7), co-segregation with disease in multiple affected family members (n=6), the variant being *de novo* (n=4), the variant being recently reported as pathogenic in ClinVar (n=2), and the finding of a different variant that results in the same amino acid change being recently reported as pathogenic in ClinVar (n=1) (**Figure 1b**). 47 VUS were downgraded to B/LB variants, but these were not further evaluated since they do not reflect a clinically actionable change. An overview of variant reclassification frequency is provided in **Figure 2a**.

**Figure 2.**
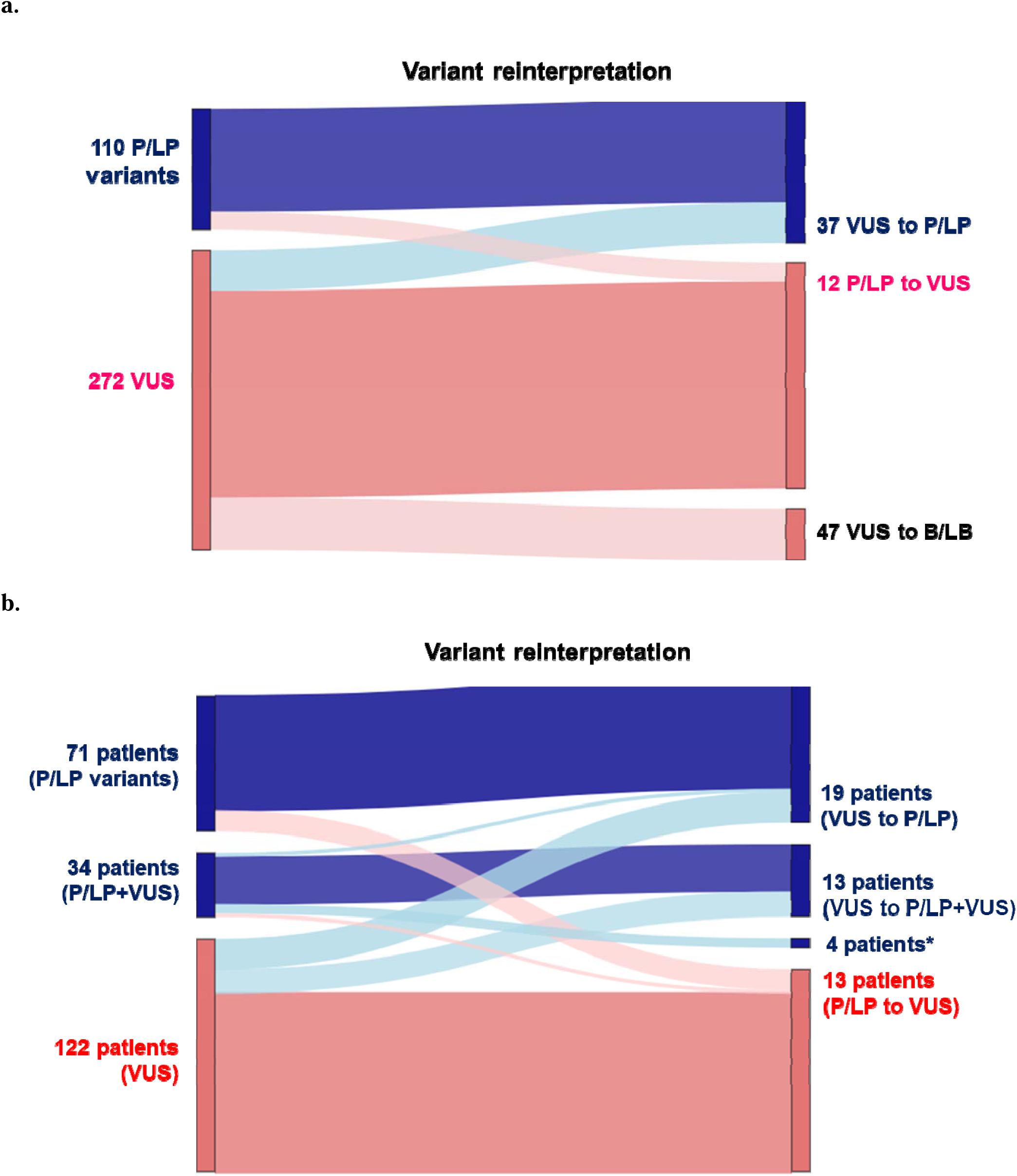
Variant and patient level reclassification: (**a**) ***Variant level reclassification***: Of 110 P/LP variants, 12 were downgraded to VUS (10.9%). Of 272 VUS, 37 were upgraded to P/LP (13.6%). In addition, 47 VUS were downgraded to B/LB. (**b**) ***Patient level reclassification***: Of 71 patients harboring only P/LP variants, 11 patients were downgraded to having only VUS. Of 122 patients harboring only VUS, 16 patients were upgraded to having only P/LP variants and 13 patients were upgraded to P/LP+VUS. Of 34 patients harboring both P/LP+VUS, 2 patients were downgraded to only VUS, 3 patients were upgraded to only P/LP variants and *4 patients had a change in the gene harboring a P/LP variant. B/LB, Benign/Likely benign; P/LP, Pathogenic/Likely pathogenic; VUS, variant of unceratin significance.

**Table 3.** VUS reclassified to pathogenic or likely pathogenic.

| Participant ID | Cardiomyopathy phenotype | Year of clinical test | Affected Gene | CDS Change | Amino Acid Change | Current ACMG criteria | Final variant classification | ACMG PS4 criteria (supporting, moderate, strong, not met) |
| --- | --- | --- | --- | --- | --- | --- | --- | --- |
| 207 | LVNC/DCM | 2014 | *ACTC1* | c.809G>A | p.Gly270Asp | PS2, PM1, PM2, PP3 (moderate) | Pathogenic | NA |
| 150 | DCM | 2018 | *ACTN2* | c.784-2A>G | NA | PVS1, PM2 | Likely pathogenic* | NA |
| 167 | DCM | Unknown | *ACTN2* | c.36C>G | p.Tyr12* | PVS1, PM2 | Likely pathogenic | NA |
| 87 | DCM | 2012 | *LMNA* | c.868G>A | p.Glu290Lys | PS2, PM2, PP3 (moderate) | Likely pathogenic† | Strong |
| 213 | HCM | 2007 | *MYBPC3* | c.442G>A | p.Gly148Arg | PM2, PP1 (strong), PP5 | Likely pathogenic*<br>† | Not met |
| 217 | HCM | 2010 | *MYBPC3* | c.1828G>A | p.Asp610Asn | PS3, PM1, PM2, PM5, PP3 (supporting) | Pathogenic | Strong |
| 31 | DCM | 2011 | *MYH7* | c.2171T>A | p.Ile724Asn | PS1, PM1, PM2, PP3 (moderate), PP5 | Pathogenic | NA |
| 57 | DCM | 2016 | *MYH7* | c.5158A>G | p.Asn1720Asp | PM1, PM2, PP3 (moderate) | Likely pathogenic | NA |
| 115 | DCM | 2019 | *MYH7* | c.3667G>A | p.Glu1223Lys | PM1, PM2, PM5, PP3 (supporting) | Likely pathogenic* | NA |
| 126 | DCM | 2019 | *MYH7* | c.4276G>A | p.Glu1426Lys | PS3, PM2, PP3 (moderate), PP5 | Likely pathogenic* | NA |
| 197 | DCM | 2022 | *MYH7* | c.1423C>A | p.Gln475Lys | PM1, PM2, PM5, PP3 (supporting) | Likely pathogenic | NA |
| 222 | DCM | 2009 | *MYH7* | c.2683C>G | p.Gln895Glu | PM1, PM2, PP1 (strong) | Likely pathogenic* | NA |
| 84 | HCM | 2017 | *MYH7* | c.4136C>A | p.Ala1379Asp | PM1, PM2, PP1 (supporting), PP3 (supporting) | Likely pathogenic | Strong |
| 151 | HCM | Unknown | *MYH7* | c.530C>T | p.Thr177Ile | PM1, PM2, PP3 (moderate) | Likely pathogenic | Moderate |
| 205 | HCM | 2006 | *MYH7* | c.1759G>A | p.Asp587Asn | PM1, PM2, PM5 | Likely pathogenic*<br>† | Strong |
| 221 | HCM | 2010 | *MYH7* | c.1220G>T | p.Gly407Val | PM1, PM2, PM5, PP3 (supporting), PP5 | Likely pathogenic | Strong |
| 179 | LVNC | 2021 | *MYH7* | c.1118C>A | p.Ala373Glu | PM1, PM2, PM5, PP3 (supporting) | Likely pathogenic | NA |
| 202 | LVNC | 2015 | *MYH7* | c.532G>A | p.Gly178Arg | PM1, PM2, PP1 (moderate), PP3 (moderate) | Likely pathogenic | NA |
| 79 | LVNC/DCM | 2015 | *MYH7* | c.1180G>A | p.Asp394Asn | PM1, PM2, PM5 | Likely pathogenic | NA |
| 126 | DCM | 2019 | *NEXN* | c.1473+1G>T | NA | PVS1, PM2 | Likely pathogenic* | NA |
| 210 | DCM | 2015 | *NEXN* | c.373C>T | p.Arg125* | PVS1, PM2 | Likely pathogenic | NA |
| 209 | LVNC/DCM | 2021 | *NEXN* | c.201G>A | p.Trp67* | PVS1, PM2 | Likely pathogenic | NA |
| 54 | HCM | 2015 | *PLN* | c.2T>C | p.Met1 | PVS1, PM2 | Likely pathogenic | NA |
| 104 | DCM | 2018 | *TNNC1* | c.376G>A | p.Glu126Lys | PS2, PM2 | Likely pathogenic | Strong |
| 5 | RCM | 2011 | *TNNC1* | c.23C>T | p.Ala8Val | PS3, PM2, PP3 (supporting), PP5 | Pathogenic | Supporting |
| 117 | DCM | 2008 | *TNNI3* | c.550G>A | p.Glu184Lys | PS2, PM1, PM2 | Likely pathogenic | NA |
| 76 | RCM/HCM | 2013 | *TNNI3* | c.484C>T | p.Arg162Trp | PM1, PM2, PM5, PP5 | Likely pathogenic | Moderate |
| 14 | DCM | Unknown | *TNNT2* | c.481C>T | p.Arg161Cys | PM1, PM2, PP3 (moderate) | Likely pathogenic | NA |
| 89 | DCM | 2016 | *TNNT2* | c.552G>T | p.Lys184Asn | PM1, PM2, PP1 (supporting), PP3 (supporting) | Likely pathogenic | NA |
| 137 | DCM | 2020 | *TNNT2* | c.316G>A | p.Glu106Lys | PS3, PM1, PM2, PP3 (moderate), PP5 | Pathogenic | NA |
| 215 | LVNC/DCM | 2014 | *TNNT2* | c.862C>T | p.Arg288Cys | PS3, PM1, PM5, PP5 | Likely pathogenic | NA |
| 28 | DCM | 2009 | *TPM1* | c.45G>T | p.Lys15Asn | PS3, PM1, PM2, PP2, PP3 (moderate) | Likely pathogenic | NA |
| 151 | HCM | Unknown | *TPM1* | c.413A>G | p.Glu138Gly | PM2, PM5, PP2, PP3 (strong) | Likely pathogenic | NA |
| 161 | HCM | 2020 | *TPM1* | c.560A>T | p.Glu187Val | PM1, PM2, PM5, PP2, PP3 (moderate) | Likely pathogenic | NA |
| 106 | DCM | 2012 | *TTN* | c.16353C>G | p.Tyr5451* | PVS1, PM2, PP1 (supporting) | Likely pathogenic | NA |
| 101 | DCM | 2010 | *VCL* | c.2949del | p.Gln971Profs* | PVS1, PM2 | Likely pathogenic | NA |
| 122 | DCM | 2018 | *VCL* | c.313C>T | p.Arg105* | PVS1, PM2 | Likely pathogenic | Strong |

\* Seven variants were reclassified by the clinical laboratory
*†3 variants were reported as P/LP in one family and as VUS in another family by the clinical testing laboratories.*
*ACM; arrhythmogenic cardiomyopathy, ACMG; American College of Medical Genetics and Genomics, CDS; coding sequence, DCM; dilated cardiomyopathy, HCM; hypertrophic cardiomyopathy, LVNC; left ventricular non-compaction, NA; Not applicable, RCM; restrictive cardiomyopathy, VUS; variant of uncertain significance.*

#### Variant burden analysis

Among the 37 variants that were upgraded from VUS to P/LP **(Supplementary Table 3)**, 11 distinct variants (in 27 cases) were identified across several extension cardiomyopathy cohorts (n=4,547) **(Table 2)**. Burden analysis confirmed 10 out of the 37 reclassified variants also met ACMG PS4 criterion for higher burden in cases compared to controls (gnomAD v4.1.0, exome dataset) (**Table 2**). The burden analysis yielded strong evidence for six variants, moderate evidence for three variants, and supporting evidence for one variant. Variants that were reclassified from P/LP to VUS were not included in the burden analysis, as the majority were downgraded due to high population allele frequency.

#### Patient level reclassification

Overall, the variant reclassifications affected 49 of 227 patients (21.6%) (**Figure 2b**). 15.5% of patients with only P/LP variants had one or more variants downgraded to a VUS. 23.8% of patients with only VUS had one or more variants upgraded to P/LP. 26.5% of patients with a combination of P/LP variants and VUS had either a downgrade of their P/LP variants and/or upgrade of their VUS in the same patient resulting in a change in affected gene.

Of note, only 8 reclassified variants (*MYBPC3* c.3628-41_3628-17del, *MYH7* c.1759G>A, p.Asp587Asn, *MYH7* c.3667G>A p.Glu1223Lys, *MYH7* c.4276G>A p.Glu1426Lys, *MYH7* c.2683C>G p.Gln895Glu, *NEXN* c.1473+1G>T, *ACTN2* c.784-2A>G and *MYBPC3* c.442G>A, p.Gly148Arg) in 12 patients had been previously reported back by the clinical genetic testing laboratory at the time of our study. Among these variants, the *MYH7* c.3667G>A p.Glu1223Lys reclassification was initiated by the clinical laboratory, and *ACTN2* c.784-2A>G reclassification was prompted by the patient’s clinical team. For the remaining variants, the reasons triggering reclassification were not available.

### Frequency of variant reclassification by subgroups

The year of clinical testing was known for 358 variants. In this group, frequency of variant reclassification did not vary by year of clinical testing i.e. before or after 2015 (**Figure 3a and 3b).** Additionally, there were no significant differences in the frequency of variant reclassification based on cardiomyopathy phenotype (HCM vs. DCM), ancestry (European vs. non-European descent), or sex (male vs. female) (all p > 0.05; **Figure 4)**. Variants identified using single gene/single targeted panel testing however showed a higher likelihood of being downgraded from P/LP to VUS compared to variants identified using multiple panels or exome sequencing (p = 0.01).

**Figure 3.**
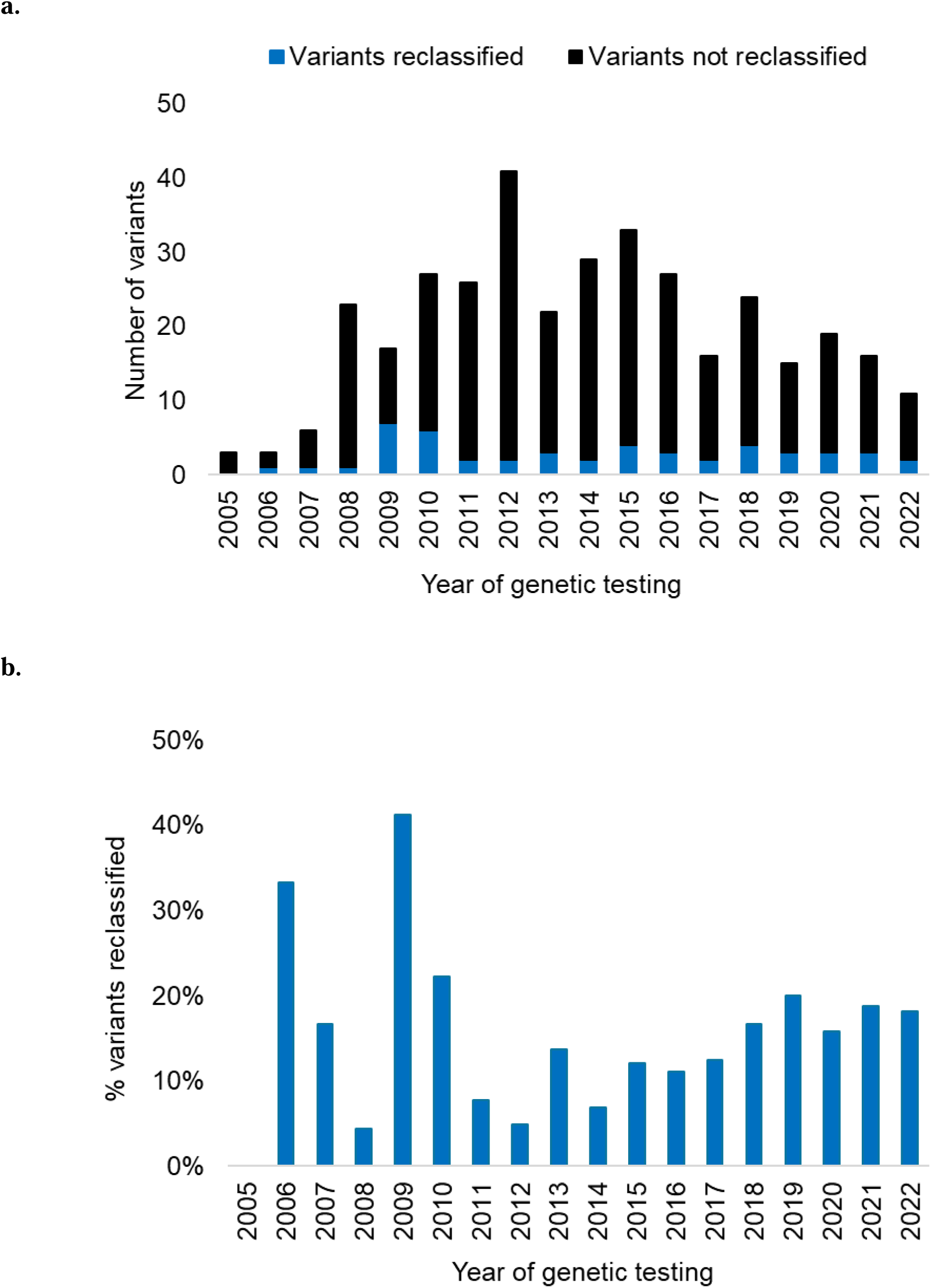
Variant reclassification by year of clinical testing. The graphs show the **(a)** frequency of variants reclassified by year of clinical testing (2005-2022); **(b)** percentage of variants reclassified by year of clinical testing. Year of testing was known for 358 variants. The average annual reclassification rate was 15%.

**Figure 4.**
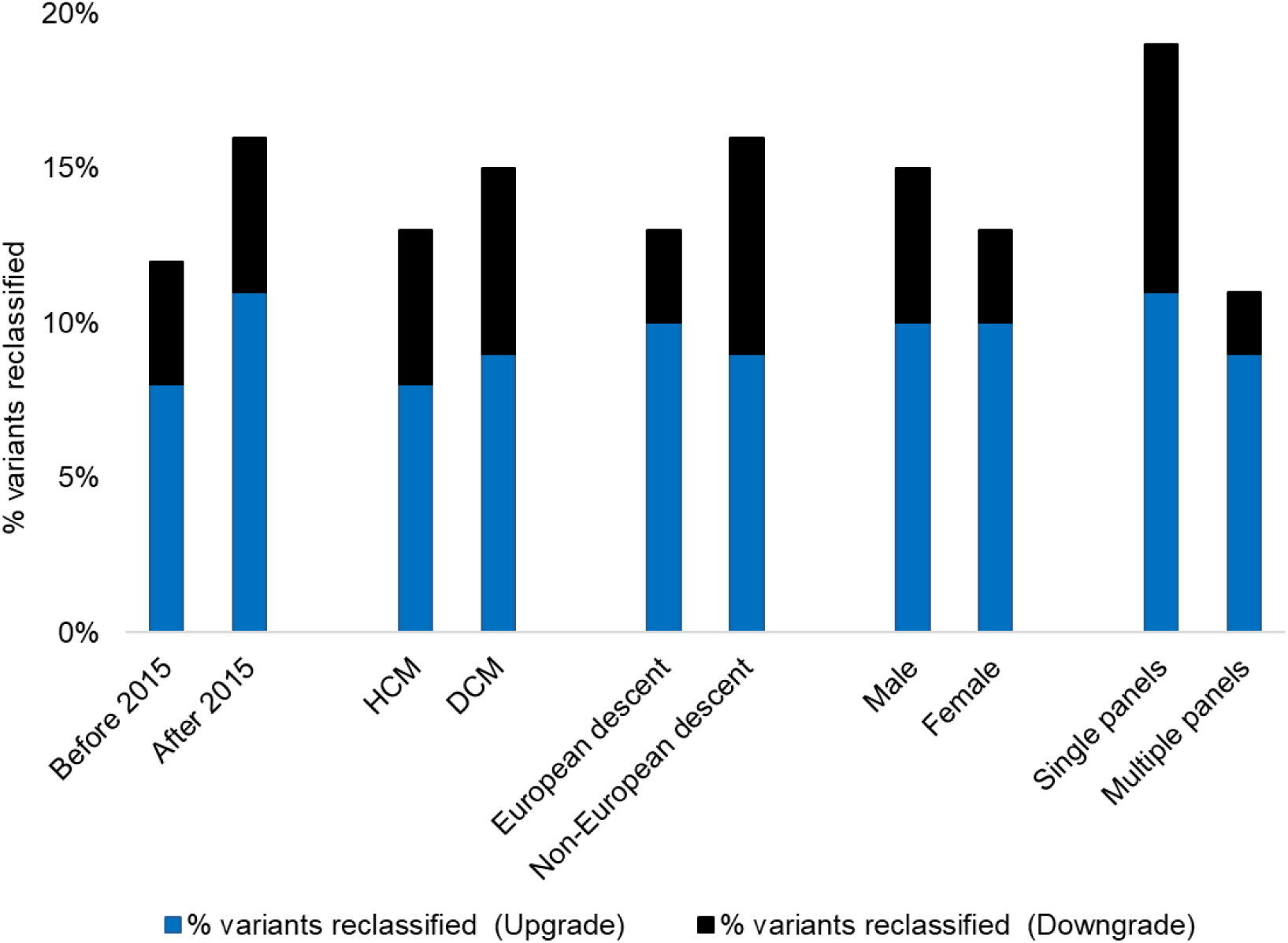
Proportion of variants reclassified by year of clinical testing, cardiomyopathy phenotype, ancestry, and sex. Proportion of variants reclassified did not differ by year of testing (before and after 2015), cardiomyopathy phenotype (HCM vs DCM), ancestry (European vs non-European descent), and sex (males vs females) (p > 0.05). Variants identified using single gene/targeted panels showed a higher frequency of P/LP variants being downgraded to VUS compared to variants identified using multiple panels or exome sequencing (p = 0.01). Blue bar = % of VUS upgraded to P/LP; black bar = % of P/LP variants downgraded to VUS. DCM, dilated cardiomyopathy; HCM, hypertrophic cardiomyopathy

## DISCUSSION

We conducted a systematic reinterpretation of variants identified on clinical genetic testing in a real-world cohort of 227 pediatric cardiomyopathy patients. The frequency of clinically actionable variant reclassification was 12.8% affecting 21.6% patients. This included downgrading of 10.9% P/LP variants to VUS and upgrading of 13.6% VUS to P/LP variants. Previous pediatric studies have reported a rate of variant reclassification ranging from 8.9%-10.3% in 71-128 patients [9, 10]. Our study with a larger cohort size reveals a somewhat higher reclassification rate and reinforces the importance of routine reinterpretation of variants on follow-up.

A detailed description of evaluation of variant pathogenicity is important to guide confidence in pathogenicity predictions [27]. In our study, we examined the criteria predominantly affecting the change in pathogenicity of the variants based on the ACMG evaluation methodology. The most common criterion for reclassification of a variant from P/LP to VUS was a higher-than-expected population allele frequency. The increase in genome sequencing including those of previously under-represented populations has enabled more accurate assessment of variant frequencies in the general population [28, 29]. Further, population-specific databases are increasingly being utilized [30], which continues to improve the accuracy of variant frequency assessments across different ancestries and will continue to enhance the precision of pathogenicity evaluation with time. The most common criterion for reclassification of a variant from VUS to P/LP was location of the variant in a mutational hotspot for the gene. Often the evaluation of a hotspot involves determining whether a variant is located within a constitutive domain of the protein or assessing the role of the domain in which the variant is located, based on literature and experimental data. In this study, reclassification was carried out using the hotspots defined by DiscoVari, the Cardiomyopathy Variant Curation Expert Panel from ClinGen, and published literature where hotspots had been identified [18, 19]. The number of genes with defined hotspots is still limited, and future efforts may enable the accurate assessment of variant hotspots in additional genes. Segregation analysis of the variant in family members and re-evaluation of inheritance contributed to a change in pathogenicity in 10 out of 81 variants (12.3%). This constitutes a substantial proportion, and as recommended in the ACMG statement [8], reclassification should include a reassessment of familial phenotype and variant inheritance on follow-up.

An important strength of our analysis was the ability to leverage several additional independent and diverse cohorts with detailed clinical annotation not routinely available in ClinVar and other public databases that helped us confirm that several of the reclassified VUS were in fact enriched in cases compared to controls on burden analysis (PS4 criterion). Therefore, it is crucial to flag variants with the potential for reclassification and to document the criteria that have the potential to change over time. It is equally important to revisit not just variant pathogenicity but also the availability of updated gene testing panels that may capture disease-associated genes that were not captured at the time of previous genetic testing. The ClinGen curation of the literature and new evidence serve an important role in informing the size and nature of gene panels.

In our study, the *MYBPC3* c.3628-41_3628-17del (Δ25bp) intronic deletion was reclassified from pathogenic to a VUS. This 25-base pair deletion is located within intron 32 of the *MYBPC3* gene and is observed in 4% to 8% of individuals of South Asian ancestry [31, 32]. Given its allele frequency which exceeds the incidence of pediatric cardiomyopathy, this variant was reclassified to a VUS (BS1 criterion). It is now known that this variant alone without *MYBPC3* c.1224-52G>A as part of a haplotype is not sufficient to produce a phenotype [33], although in the adult population, it has been associated with myocardial diastolic dysfunction [34].

There were no significant differences in the proportion of patients reclassified before and after 2015, when the ACMG/AMP guidelines were first published. Of note, 2 of 11 variants (18%) identified on clinical testing in 2022, the most recent year included, showed a change in pathogenicity within 2 years of clinical reporting. The downgrading of *RRAGD* c.289_303del p.Thr97_Cys101del from LP to VUS was based on the application of in silico tools (PP3), which indicated that the variant was not deleterious. Conversely, the upgrading from VUS to LP of *MYH7* c.1423C>A p.Gln475Lys was because of evidence of a mutational hotspot at the codon (PM1) and a differential missense change at the same codon (*MYH7* c.1425G>T p.Gln475His), which had been previously reported as pathogenic (PM5) in 2022, further supported by in silico analysis using AlphaMissense issued in 2023 (PP3). Hence, it is important to reevaluate variants every few years due to rapid advances in genomic characterization of cases and controls. In this regard, there remains a gap in regular reinterpretation of genetic variants by testing labs. Only 8 of the 49 variants reclassified by us were reported back to the clinical team by the original testing laboratory. The ACMG and the other reports recommend that clinical laboratories reevaluate variants at least every three to five years and the costs of recontacting physicians can be built into original testing costs [8, 35, 36]. The Canadian College of Medical Geneticists recommends that laboratories reanalyze variants when a healthcare provider initiates the request [37]. This is important because laboratories do not always have access to updated family phenotype information. This emphasizes that periodic reinterpretation should, where feasible, be built into the ongoing clinical care of patients by their healthcare providers. The ACMG in its most recent policy statement emphasizes that “re-contact is a shared responsibility” [38], indicating that both clinicians and clinical laboratories should take on an increasing role in assessing and communicating variant reinterpretations to patients. In addition, patients and families should be advised to notify their physician if family genotype or phenotype information changes on follow-up.

The present analysis revealed that variants identified using single gene / single targeted panel showed a significantly higher reclassification rate compared to multiple panels, primarily driven by a greater proportion of variants that were downgraded from P/LP to VUS. Given the high degree of genetic heterogeneity with overlap in genetic etiology between different cardiomyopathies, this finding suggests that comprehensive genetic testing is more likely to identify true P/LP variants with fewer false-positives compared to targeted gene panels. In the future, it may be useful to consider developing a framework that considers the type of genetic test performed when determining which cases should be prioritized for variant reinterpretation.

It is important to acknowledge as a limitation that we were not privy to the exact rationale and criteria for the original variant classification used by the clinical laboratory (especially for ACMG criteria that were only published in 2015) or their threshold for recontacting physicians with updated reclassifications.

### Clinical significance

The findings of a meaningful change in variant pathogenicity in 21.6% patients has clinical implications. The 14 patients in whom P/LP variants were downgraded will require all previously genotype-negative family members to return for ongoing clinical screening. For the 35 patients in whom VUS were upgraded to P/LP, family members can now be offered cascade genetic testing to determine who is at risk and needs ongoing clinical surveillance and who is genotype-negative and can be discharged from follow-up. Providing timely feedback to patients regarding changes in variant classification can lead to changes in follow-up protocols and may improve prognosis for other family members. A re-evaluation system for a broad range of variants, including those that emerge as children transition into adulthood is essential given the long-term prognosis of pediatric patients.

## Conclusions

One in 5 patients with childhood onset cardiomyopathy had a clinically actionable change in variant pathogenicity on follow-up that impacts clinical and genetic family screening recommendations. This highlights the importance of serial reevaluation of variant pathogenicity by clinical providers as well as by clinical genetic testing laboratories.

## ACKNOWLEDGEMENTS

We acknowledge the patients and families participating in the Labatt Family Heart Centre Biobank at the Hospital for Sick Children for access to study data. Part of this research was made possible through access to the data generated by the 2025 French Genomic Medicine Initiative.

## ETHICS APPROVAL

The study was approved by the Institutional Research Ethics Board of The Hospital for Sick Children. Written informed consent to participate was obtained from all patients and/or their parents/legal guardians. The study protocol adhered to the Declaration of Helsinki.

## DATA AVAILABILITY

De-identified data analyzed in this study are available in the main and supplemental tables, and additional data are available from the corresponding author on reasonable request.

## AUTHOR CONTRIBUTIONS

TS, RL, and SM conceptualized and planned the study, performed data analysis, drafted the initial manuscript, and made significant revisions. RA collected the clinical data. PX, LZ and RJ assisted with bioinformatics and clinical data analysis. CS, EMB, KH, NRB, EAT, PSD, VJR, JI, and NH contributed variant data interpretation from their respective cardiomyopathy cohorts. SM secured funding for the project. All authors approved the final manuscript as submitted and agree to be accountable for all aspects of the work.

## Abbreviations

ACM: Arrhythmogenic ventricular cardiomyopathic
AMP: The American College of Medical Genetics and Genomics/Association for Molecular Pathology
B/LB: Benign/Likely benign
CI: Confidence interval
DCM: Dilated cardiomyopathy
HCM: Hypertrophic cardiomyopathy
LVNC: Left ventricular noncompaction cardiomyopathy
OR: Odds ratio
pLI: probability of loss-of-function intolerant
P/LP: Pathogenic/Likely pathogenic
RCM: Restrictive cardiomyopathy
VUS: Variant of uncertain significance

## SUPPLEMENTARY MATERIAL

**Table S1** All cardiomyopathy variants reevaluated (n=382)

**Table S2** Reclassified VUS observed in extension cohorts

**Table S3** Burden analysis for upgraded VUS in extension cohorts

